# Comparing organ donation decisions for next-of-kin versus the self: Results of a national survey

**DOI:** 10.1101/2021.07.12.21260242

**Authors:** Christopher W. Liu, Lynn N. Chen, Amalina Anwar, Boyu Lu Zhao, Clin K. Y. Lai, Wei Heng Ng, Thangavelautham Suhitharan, Vui Kian Ho, Jean C. J. Liu

## Abstract

**Objectives:** Intensive care audits point to family refusal as a major barrier to organ donation. In this study, we sought to understand refusal by accounting for the decision-maker’s mindset. This focused on: (1) how decisions compare when made on behalf of a relative (versus the self); and (2) confidence in decisions made for family members.

**Design:** Cross-sectional survey in Singapore.

**Setting:** Participants were recruited from community settings via door-to-door sampling and community eateries.

**Participants:** 973 adults who qualified as organ donors in Singapore.

**Results:** Although 68.1% of participants were willing to donate their own organs, only 51.8% were willing to donate a relative’s. Using machine learning, we found that consistency was predicted by: (i) religion, and (ii) fears about organ donation. Conversely, participants who were willing to donate their own organs but not their relative’s were less driven by these factors, and may instead have resorted to heuristics in decision-making. Finally, we observed how individuals were overconfident in their decision-making abilities: although 78% had never discussed organ donation with their relatives, the large majority expressed high confidence that they would respect their relatives’ wishes upon death.

**Conclusions:** These findings underscore the distinct psychological processes involved when donation decisions are made for family members. Amidst a global shortage of organ donors, addressing the decision-maker’s mindset (e.g., overconfidence, the use of heuristics) may be key to actualizing potential donors identified in intensive care units.

**Strengths and Limitations of this Study:**

- We used a multi-disciplinary approach combining psychology theory and machine learning analyses to understand family refusal in a novel manner.
- We directly compared organ donation decisions made from the self versus for next-of-kin, and also documented overconfidence in the decision-making process.
- The study was conducted in an urban setting and may not apply to rural contexts.

## Introduction

Solid organ transplantation is widely recognized as a life-saving treatment for end-stage organ failure.^1^ Each year, nearly 150,000 organs are transplanted worldwide, improving patient outcomes and the quality of life.^2^ Nonetheless, the number of transplant surgeries performed each year caters to a mere 10% of the demand, with numbers constrained by a global shortage of donor organs.^2^

To address the shortage, there have been concerted efforts to increase the rate of deceased organ donation worldwide.^3,4^ In terms of bottlenecks, audits of intensive care units implicate family refusal as a major barrier to donation.^5,6^ Often, when a deceased patient is identified as a potential donor, family members are consulted prior to organ retrieval. In an average of 2 in 5 of these cases, however, the family refuses to donate the patient’s organs.^5,7^

### Accounting for Family Refusal

A large body of research has sought to account for family refusal by reviewing medical records or interviewing next-of-kin.^7-10^ One crucial predictor is whether relatives are aware of the deceased’s wishes: in general, next-of-kin enact a patient’s choice if they are aware of it.^7,11^ However, an estimated 50% of families do not know the deceased’s preferences, and are influenced instead by factors ranging from the context of the request (e.g., timing, background of the requestor), to pre-existing characteristics of the decision-maker (e.g., demographics, beliefs about death, views on organ donation).^7-9,12,13^ By and large, these factors overlap greatly with those predicting donation choices for the self.^9,14,15^

Although this overlap may suggest similarity in the self-versus family-decision making process, it cannot explain the well-documented discrepancy between: (1) high public endorsement of organ donation (for the self), versus (2) low rates of family acceptance (when deciding for next-of-kin).^16,17^ Likewise, when participants are surveyed, they report being more willing to donate their organs than a relative’s organs^18,19^ – a stark drop-off not observed for other death-related procedures (e.g., autopsy).^18^ Together, these systematic shifts suggest that the psychology of decision-making may differ when it comes to next-of-kin. This is a piece of the organ donation puzzle that we know little about, but may be central to actualizing potential donors identified in intensive care units.

### Decision Making for the Self Versus for Others

Outside the field of organ donation, social scientists have described how choices change when they are made on behalf of others.^20^ For example, risks may be assessed differently because third party decision-makers are detached, perceive the situation differently, or – conversely – feel a greater weight of responsibility for the outcomes (leading to more conservative choices).^20^ Pertaining to medical decision-making, however, these general tendencies fall short of ethical guidelines prescribing how decisions *should* be made for incapacitated patients.^21^

As a case in point, ethicists typically argue for ‘substituted judgment’: if a patient’s wishes have not been documented, a surrogate family member should strive to make the same decision that the patient would have made for him-or herself.^21^ However, extant research suggests that this rarely occurs. When decision-makers are shown hypothetical end-of-life scenarios and asked to predict their family member’s treatment preferences, they typically make inaccurate judgments.^22^ Nonetheless, both surrogates and patients report high confidence that the surrogate will carry out the patient’s wishes – a level of overconfidence that impedes advance care planning and conversations about end-of-life wishes.^23^ This overconfidence, in turn, results in high levels of surrogate stress at the point of decision-making.^24,25^

### The Current Study

Although surrogate decision-making has been described for end-of-life patient care, analogous research has not been conducted for organ donation. To address this gap, we used data from a large cross-sectional survey to understand the distinct mind-set of decision-makers as they contemplate organ donation for a relative.

First, we hypothesized that there would be a greater willingness to donate one’s own organs than a family member’s. Correspondingly, we sought to identify factors predicting the shift from being a willing organ donor (for the self) to refusing organ donation (for a family member). This strategy stands in contrast to the typical analysis in organ donation studies, which have focused on predicting decisions for the self or the family member independently (rather than the discrepancy between these).^10^

Second, drawing from research on surrogate decisions for incapacitated patients,^16,26^ we investigated the role of overconfidence in family refusal. Specifically, we hypothesized that overconfidence would similarly characterize the mindset of individuals making organ donation decisions on behalf of another.

## Method

### Study Design and Population

Our study was conducted in Singapore, a city state in Asia where an opt-out system is in place for donation of the kidney, liver, heart, and cornea (implemented via the country’s Human Organ Transplant Act).^27^ Additionally, an opt-in system applies to donation of all other organs and tissues (e.g., skin, bone), and for individuals who do not qualify for presumed consent.^28^ Under the opt-in context, family members can voluntarily donate relatives’ organs if no prior decision has been registered.^28^ In the opt-out context, although next-of-kin cannot legally over-ride the presumed consent once brain death has been certified, they are often consulted to identify a patient’s expressed or implied wishes prior to the impending brain or circulatory death. This may lead to withdrawal of cardiopulmonary support in light of medical futility, prior to brain death assessment.^29^

From September 2016 to March 2019, we surveyed 973 adults who qualified as organ donors under Singapore’s Human Organ Transplant Act.^27^ As the inclusion criteria, participants were: (i) citizens or permanent residents of Singapore, (ii) who were aged ≥ 21 years old. In line with the Act, participants were excluded if they were mentally incapacitated.

The National University of Singapore’s Institutional Review Board approved all procedures (IRB A-16-131), the study protocol was registered on ClinicalTrials.gov (NCT04303624), and participants gave verbal consent.

### Survey Administration

To obtain a representative sample, participants were recruited both door-to-door and via community eateries. For door-to-door recruitment, households were identified using cluster-sampling (by selecting districts in Singapore) followed by simple random sampling (by selecting postal codes within each district). For each postal code, trained interviewers then knocked on doors systematically during weekday evenings and weekends, selecting as respondent the member of the household whose birthday was most recent. (Details of this methodology have been published elsewhere.^14^) For recruitment via community eateries, we visited two popular food halls during the weekday lunch period (12-4pm). Trained interviewers then systematically approached each occupied table and invited diners to participate.

As part of a larger study, data collection took place in three waves (Wave 1: September 2016-July 2017, Wave 2: May 2017-July 2017, and Wave 3: July 2018-March 2019), and responses were merged to form a single database. Appendix 1 gives a detailed breakdown of each collection phase and how survey versions differed. (As the key results did not change as a function of wave, subsequent analyses collapsed across this factor.)

### Main Outcome Measures

Following guidelines on survey construction in healthcare,^30^ questionnaire items were developed using prior measures in both the organ donation and medical surrogacy literature. ^18,23^ The primary outcome variables were: (1) deciding on organ donation for the self (whether participants would like to donate their organs upon death), and (2) deciding on organ donation for a family member (whether participants would like to donate a family member’s organs upon death).

To examine over-confidence, a subset of participants (from Wave 2) rated how confident they were that – upon their family member’s death – they would respect his/her wishes regarding organ donation (rated using 5-point scales anchored on one end with 1=“Not confident at all” and 5=“Absolutely confident”). These were compared to participants’ self-reports of: (1) whether the family member had ever discussed his/her wishes concerning organ donation, and (2) the extent to which participants thought they were aware of their family member’s wishes (rated on a 5-point scale with 1=“Absolutely unaware” and 5=“Absolutely aware”, with ratings of 4 or 5 representing awareness). For comparison, an analogous set of questions were asked regarding decisions family members may make for the participant (in Waves 1 & 2).

### Predictor Variables

As predictors, we collected a range of demographic variables commonly used to account for individual or family decision-making: gender, age, self-identified race/ethnicity, religion, marital status, education, house type, and household size.

Additionally, participants in Wave 1 and 2 rated the extent to which each of 18 statements characterized their organ donation concerns for a family member. These questions, drawn from previous studies on the topic,^18^ asked participants about their discomfort pertaining to: (i) mortality (1 item: “I am uncomfortable thinking about my family member’s death in general”); (ii) clinical management (5 items: e.g., “I am afraid that my family member may not get optimal medical care; that the doctors may prematurely declare him/her dead just to have his/her organs donated); (iii) donation suitability and equitability (4 items: e.g., “I am concerned that my family member’s organ donation may end up benefitting rich people more than the average person”); (iv) post-death procedures (3 items: e.g., “I am uncomfortable with someone cutting up my family member’s dead body); (v) honoring the deceased (1 item: “I find organ donation disrespectful to a deceased family members”); (vi) spirituality (3 items: e.g., “I am afraid that my family member’s reincarnation may be influenced by organ donation”); and (vii) family dynamics (1 item: “I am afraid that other family members may not agree with my decision”).^18^ Each item was rated using a 5-point scale anchored on one end with 1=“Strongly disagree” and 5=“Strongly agree”, and scores were averaged to form a scale (Organ Donation Fears: Family; Cronbach’s α = 0.94). Similarly, participants rated a corresponding set of concerns for themselves (Organ Donation Fears: Self; Cronbach’s α = 0.93).

As decisions about family members may be shaped by culture, participants in Wave 1 completed the Cultural Orientation Scale, indicating whether they viewed themselves ‘collectively’ – as part of social groups such as families – or ‘individualistically’, independent of groups.^31^ Using 5-point scales (1=“Strongly disagree” and 5=“Strongly agree”), participants rated: 4 items on hierarchical relationships within a collective (Vertical Collectivism, e.g., ‘It is my duty to take care of my family, even when I have to sacrifice what I want’; Cronbach’s α = 0.80), 4 items on equal relationships within the collective (Horizontal Collectivism, e.g., ‘To me, pleasure is spending time with others’; Cronbach’s α = 0.78), 4 items on hierarchical relationships amongst individuals (Vertical Individualism, e.g., ‘Winning is everything’; Cronbach’s α = 0.74), and 4 items on equal-standing relationships amongst individuals (Horizontal Individualism, e.g., ‘I’d rather depend on myself that others’; Cronbach’s α = 0.76). Finally, participants also completed the Cultural Values Scale, rating 5 items on how power should be shared (Power Distance, e.g., “People in higher positions should make most decisions without consulting people in lower positions”; Cronbach’s α = 0.83).^32^ For each dimension, items were summed to form subscale scores.

### Data Analysis

For the primary analyses, survey responses were summarised with counts (%) and medians (with interquartile ranges [IQR]). To predict shifts in decision-making for family members relative to the self, we performed a classification tree analysis using recursive partitioning (“rpart”).^33,34^ This machine learning model allows multiple variables to be analyzed simultaneously while accommodating complex relationships between predictors.^35^ For the model, input variables were: the 8 demographic variables, scores on both organ donation fear scales (for a family member and for the self), scores on the 5 culture-related scales (Vertical Collectivism, Horizontal Collectivism, Vertical Individualism, Horizontal Individualism, and Power Distance), and overconfidence metrics for both family members and the self (whether organ donation discussions had been taken, confidence in decision-making, and family awareness). To avoid overfitting, the final tree was obtained by requiring at least 10 participants per subgroup and a minimum Gini reduction of 0.01 within subgroups.

Finally, we reported two forms of inferential statistics: chi-square tests of independence (to examine the relation between various donation and overconfidence metrics), and Tukey’s test (as a post-hoc test following recursive partitioning). For all statistical tests, Type 1 Decision Wise Error Rate was controlled at α = 0.05. All analyses were performed in SPSS 25 and R 3.6.2.

### Patient and Public Involvement

We did not involve patients or the public in the formulation of research questions or in data analysis. However, survey items were designed based on the authors’ clinical experience discussing organ donation decisions with families.

## Results

### Description of the Sample

Participants were 973 adults from the general community (Figure 1). As shown in Table 1, respondents were equally likely to be male (469 [48.2%]) or female (472 [48.5%]), and had a mean age of 40.26 years (SD: 12.57 years). Respondents were comparable to Singapore’s general population in: gender, ethnicity, marital status, and religion (≤5% difference). Notably, there was a greater representation from the 21-49 age group (64.8% vs. 54.4%), from residents of public housing (89.7% vs. 78.3%), and from recipients of tertiary education (68.0% vs. 48.2%).^36^. On the other hand, participants were less likely to be from 1-2 member households than the general population (16% vs. 37%).^36^

**Table 1.**
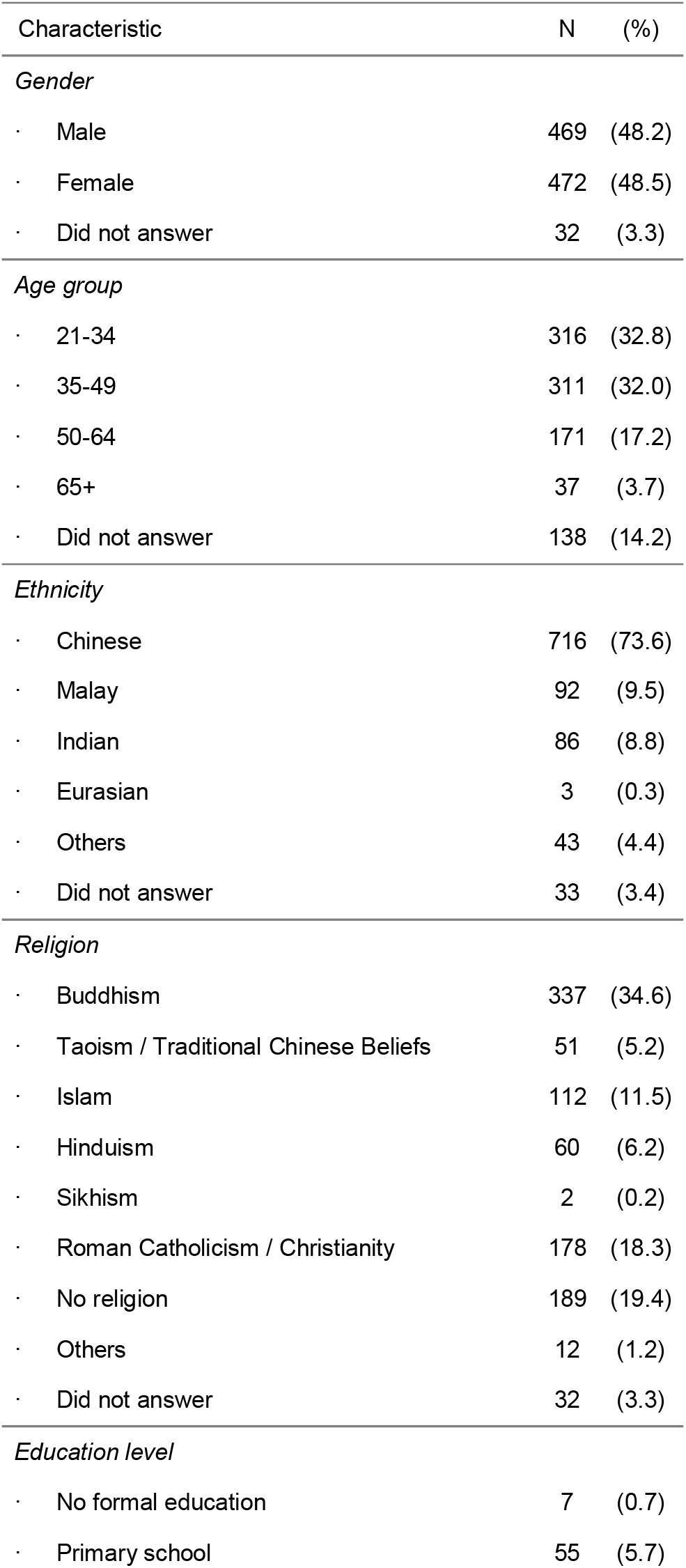
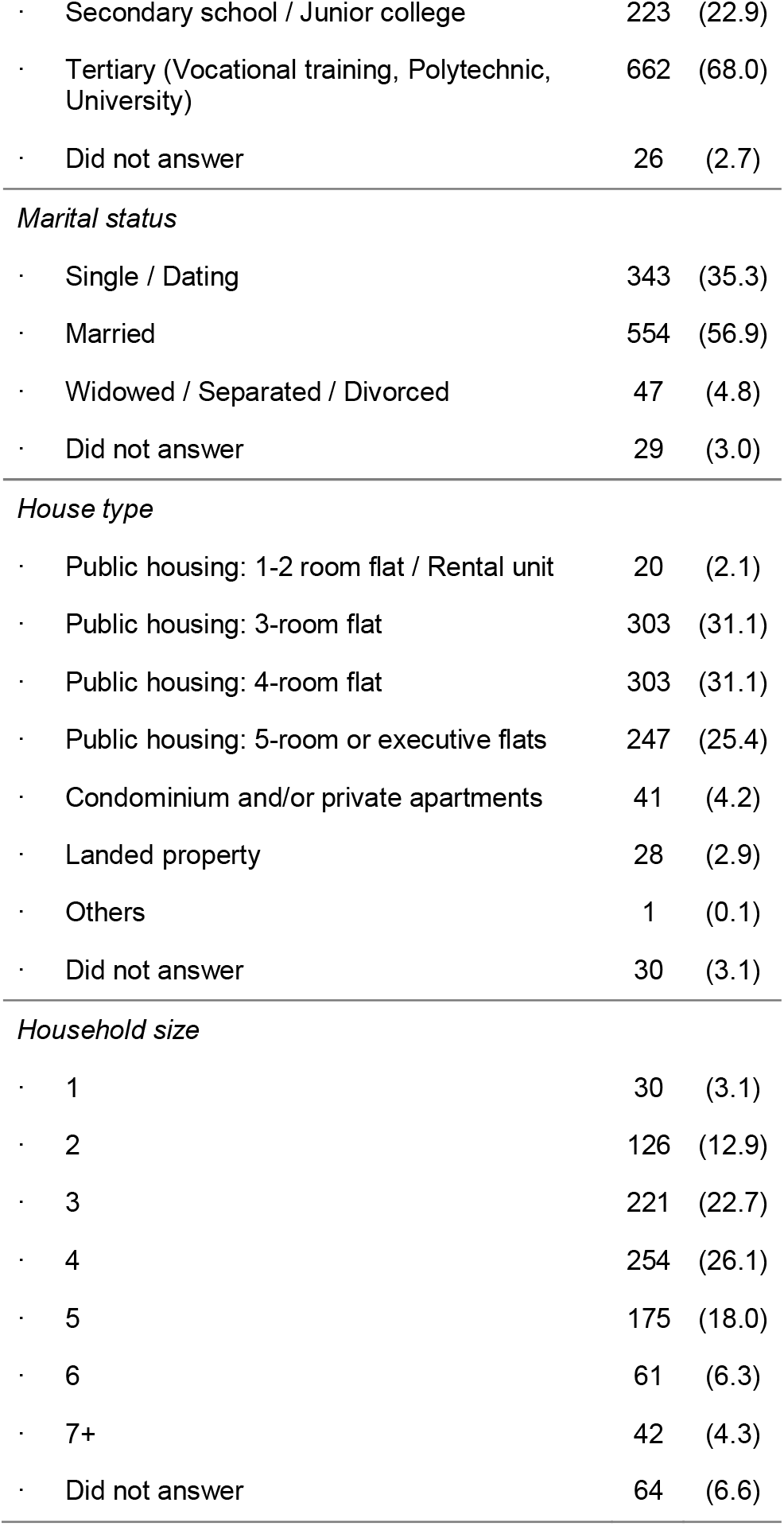
Baseline characteristics of survey respondents.

**Figure 1.**
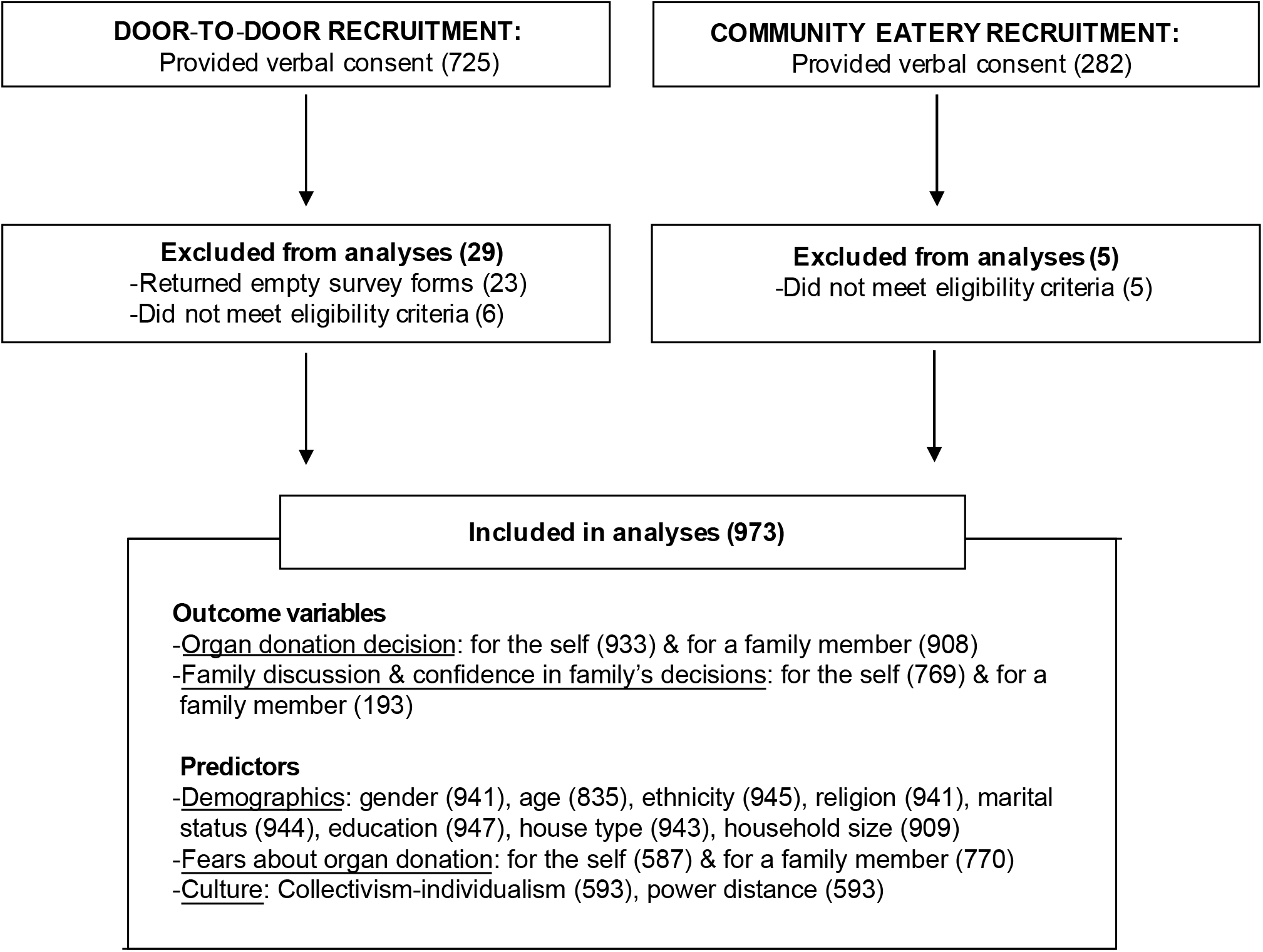
Flowchart of participant inclusion.

### Deciding for the Self versus a Family Member

As shown in Figure 2, 1 in 5 participants (22.1%, 95% CI: 19.6-25.1%) were willing to donate their own organs but were unwilling to donate a family member’s. This corresponded to a drop-off in willingness to donate, with lower rates reported when deciding for family members than for themselves (95% CI of difference: 11.7-20.9%); *X*^2^(1) = 48.29, *p* < 0.001).

**Figure 2.**
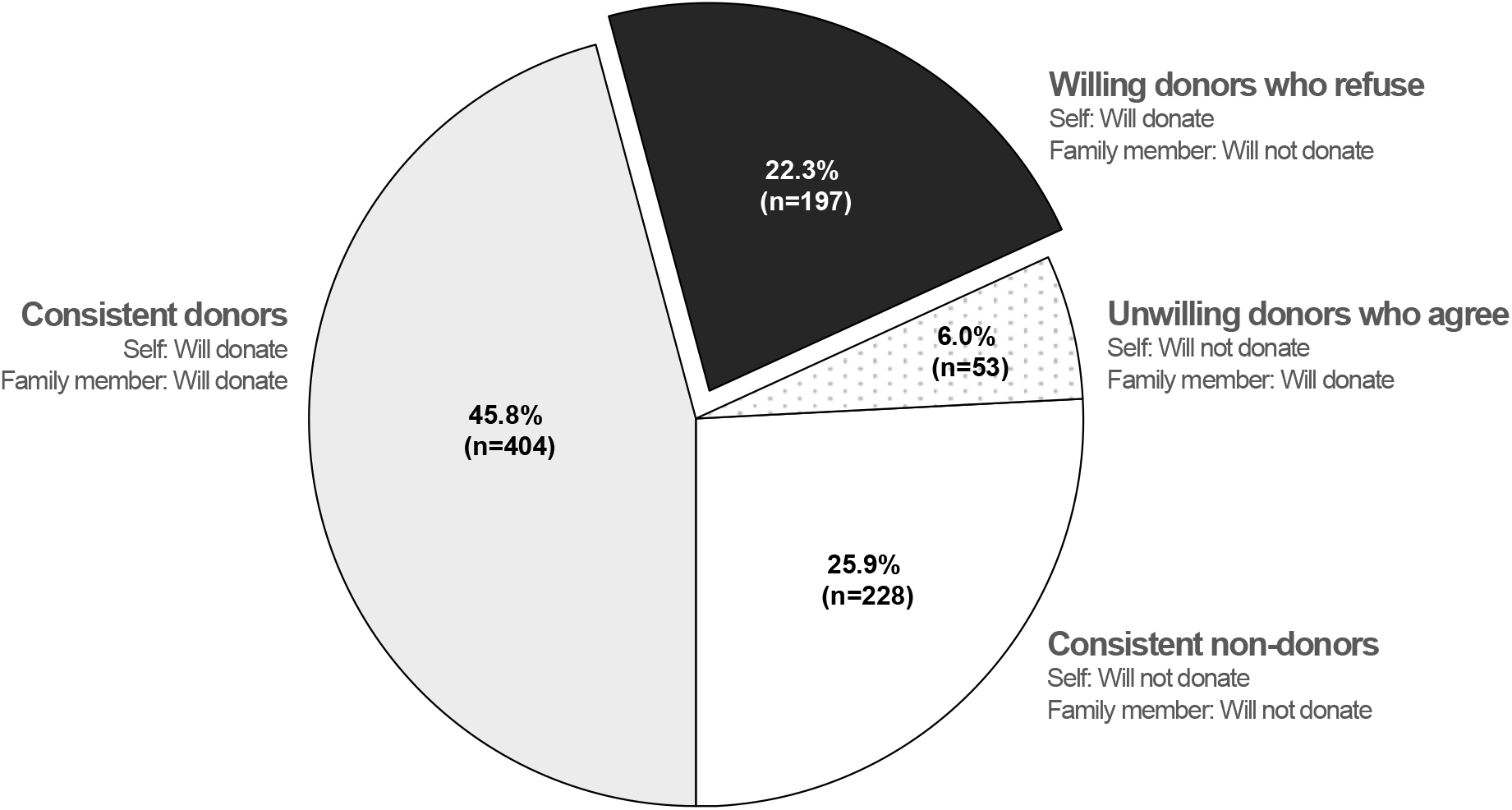
The distribution of participants as a function of their willingness to: donate their own organs, and donate a family member’s organs.

### Predicting Inconsistency in Decision-Making

To understand inconsistency in decision-making, we used recursive partitioning to predict which of four possible categories participants belonged to: (1) consistent donors (willing to donate for both the self and for family members), (2) consistent non-donors (unwilling to donate in either context), (3) unwilling donors who agree (unwilling to donate for the self, but willing to donate for family members), or – importantly – (4) willing donors who refuse (willing to donate for the self, but refusing for family). The final tree model identified participants’ organ donation fears (self and family) and religion as the key predictors of category membership (Figure 3).

**Figure 3.**
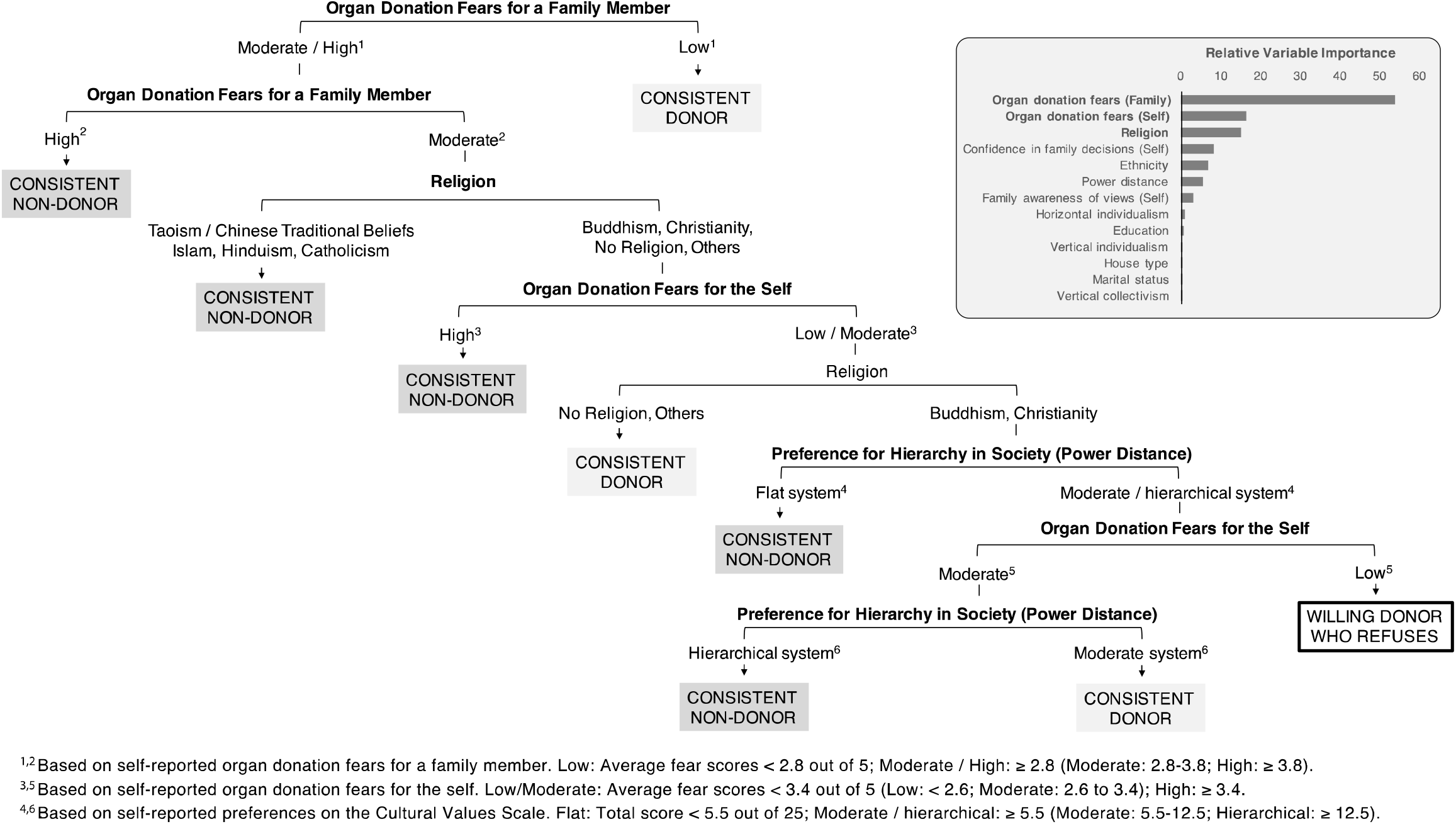
A machine learning technique – recursive partitioning – was used to predict which of 4 categories participants belonged to: (i) consistent donors to donate for both the self and for a relative); (ii) consistent non-donors (unwilling to donate in both cases); (iii) willing donors who refuse (being willing to for themselves but not for next-of-kin); or (iv) unwilling donors who agree (being unwilling for the self but willing for next-of-kin). The final model, present flow chart, shows how participant information was used to maximise information gain: at each level of the chart, a factor was chosen that allowed the most of participants to be categorised. As shown in the bar graph, participants’ organ donation fears and religion emerged as the key predictors (model class accuracy: 52%, above the chance level of 25%).

As shown in Figure 4, consistency was marked by more extreme values in organ donation fears. Consistent non-donors reported higher organ donation fears both for themselves and for family members (*p*-values for Tukey post-hoc test: all ≤0.003 for the self, and all ≤0.008 for family members), whereas consistent donors reported lower organ donation fears, relative to other groups (*p*-values for Tukey post-hoc test: all <0.001). Consistent non-donors were also more likely to report a religious affiliation (Taoism or Chinese traditional beliefs, Islam, Hinduism, or Catholicism), whereas consistent donors were more likely to have no religion.

**Figure 4.**
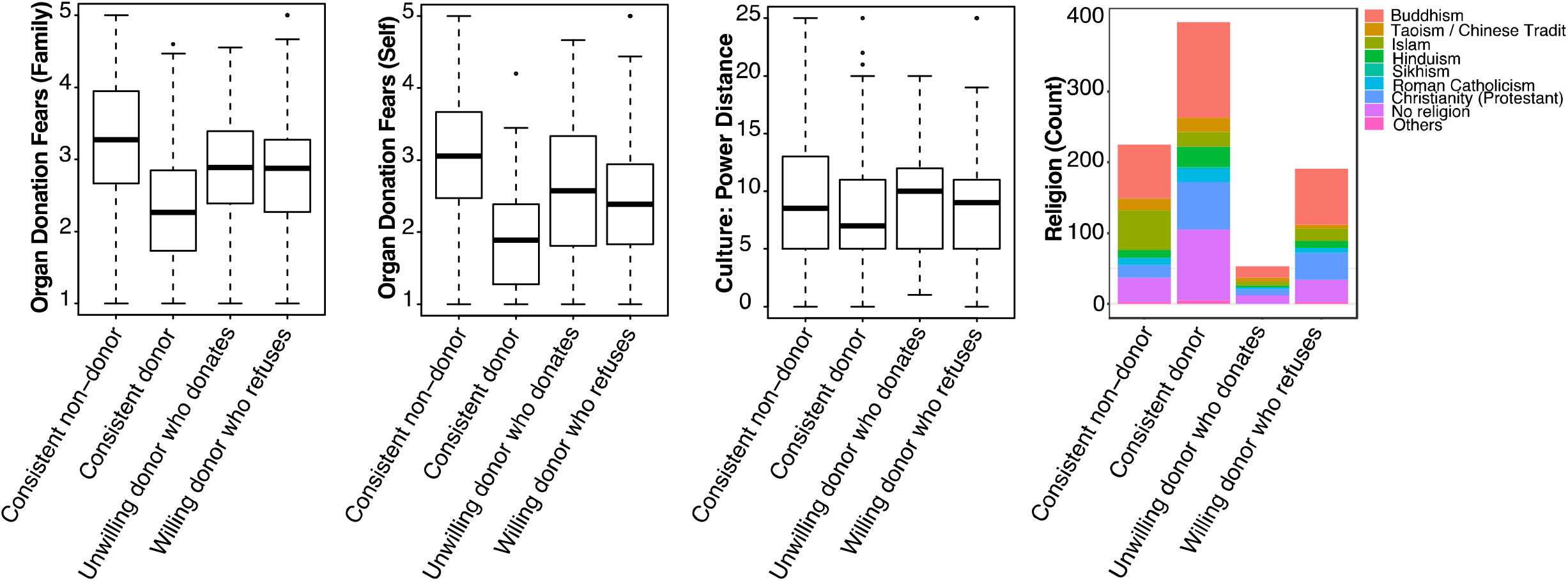
Participants’ organ donation fear scores (for a family member and for themselves), power distance scores, and religion as a function of their d decisions.

In contrast, predicting willing donors who refused family donation was not as straightforward. This group was identified as those who met a series of criteria: (i) having low organ donation fears for themselves (scores of <2.6 out of 5) but greater fears for family members (between 2.8-3.8 out of 5); (ii) having Buddhist or Christian beliefs; and (iii) not holding extreme views about how power should be shared in society (power distance scores ≥5.5). Notably, none of the criteria uniquely identified this group. For example, when paired with other criteria, Buddhism and Christianity were also associated with consistent non-donors and donors.

### Overconfidence in Family Decision-Making

Following the surrogacy literature,^23^ we examined whether overconfidence characterized family decision-making. As shown in Figure 5, nearly 4 in 5 participants (78%, 95% CI: 72.1-83.9%) had never discussed organ donation with a close family member they could be tasked to make decisions for. Correspondingly, only 1 in 3 participants (29.9%, 95% CI: 23.4-36.4%) were aware of their relative’s organ donation wishes. We note, however, that higher rates of awareness were reported amongst those who had engaged in prior discussion (80%, 95% CI: 67.6-92.4%) than those who had not (22.2%, 95% CI: 15.4-29.0%); χ2(4, *N* = 184) = 55.58, *p* < 0.001.

**Figure 5.**
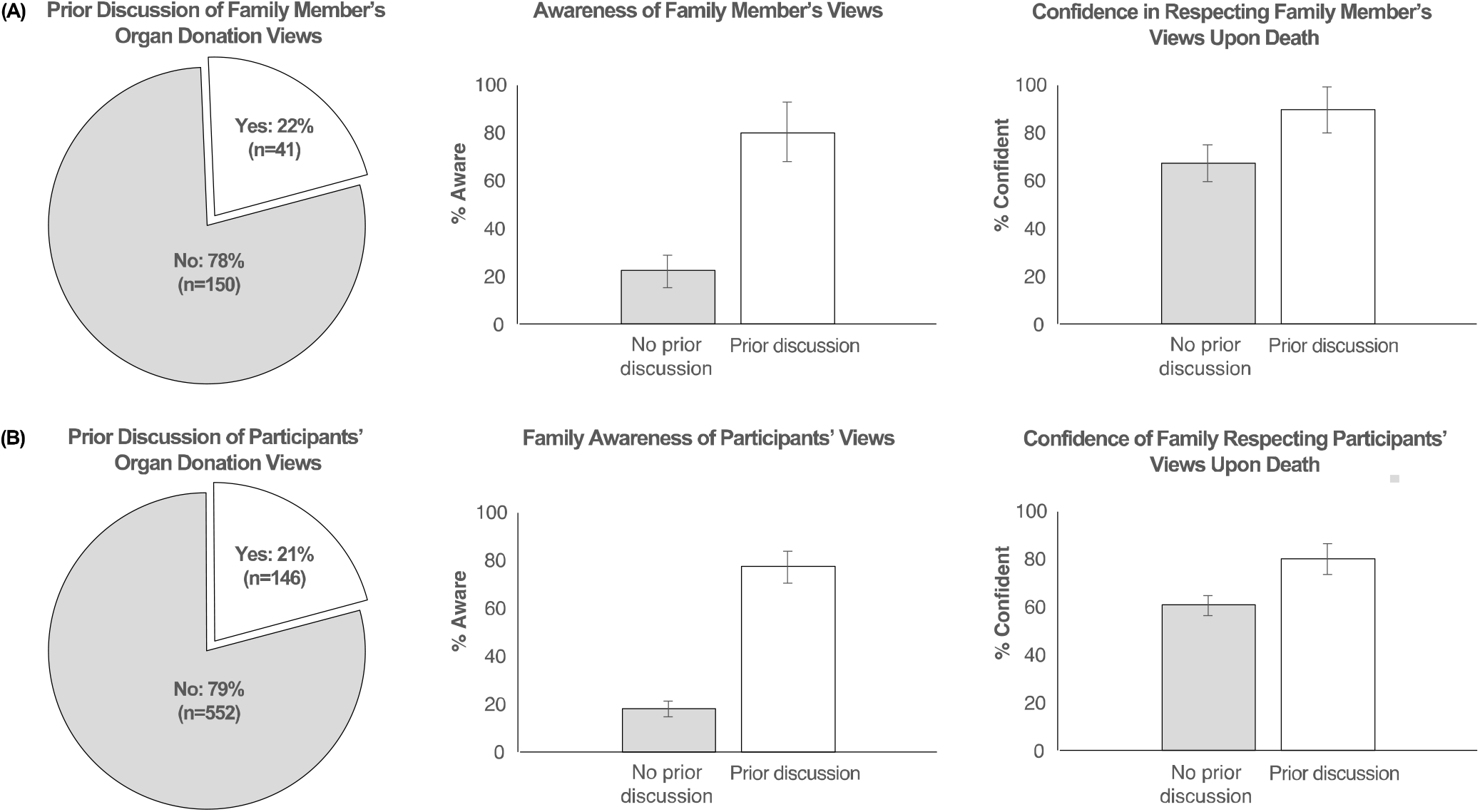
(A) Participants reported whether they had discussed organ donation with a close family member they may be tasked to make decisions for (left panel). The bar graphs depict the percent of participants in each category (no prior discussion vs. prior discussion) who reported being aware of their family members’ views (middle panel), and confident that they would respect their family members’ wishes upon death (right panel). (B) Similarly, participants reported whether they had discussed their own views with their family (left panel), whether their family was aware (middle panel), and how confident they were that their family would carry out their wishes (right panel). Vertical lines represent 95% confidence intervals.

Although few participants were aware of family members’ wishes, participants were nonetheless very confident that they would respect these wishes upon their relatives’ deaths. When asked to rate how confident they were (using a 5-point scale), the median rating was 5 – “absolutely confident” (IQR: 3 to 5). Further, confidence was high both amongst those who had previously discussed donation with their relatives (89.7% gave a rating of 4 or 5; 95% CI: 80.2-99.2%), and amongst those who had not (67.1%, 95% CI: 59.4-74.8%); confidence did not differ significantly as a function of whether participants had engaged in prior discussion (χ2(4, *N* = 182) = 8.44, *p* = 0.08).

As a comparison, we examined the corresponding questions pertaining to participants’ own wishes on organ donation (Figure 5). The results were near-identical: 4 in 5 participants (79.1%, 95% CI: 76.1-82.1%) had never discussed their wishes with their family, and only 1 in 3 (29.9%, 95% CI: 26.7-33.1%) perceived that their family was aware of their wishes. Nonetheless, the majority of participants felt confident that their family would carry out their wishes upon death (64.0% gave a rating of 4 or 5; 95% CI: 60.6-67.4%).

Finally, we ran exploratory analyses comparing this set of metrics amongst participants of the four decision categories previously identified (consistent non-donors, consistent donors, unwilling donors willing to donate relatives’ organs, and willing donors who refuse family donation). As shown in Figure 6, donation decisions related to participants’ awareness of their relatives’ wishes, χ2(3, *N* = 182) = 9.01, *p* = 0.03. Namely, consistent non-donors were more likely to report awareness than: consistent donors (χ2(1, *N* = 147) = 5.81, *p* = 0.02), unwilling donors who donate relatives’ organs (χ2(1, *N* = 25) = 3.95, *p* = 0.047), or willing donors who refuse family donation (χ2(1, *N* = 48) = 7.31, *p* = 0.007); whereas awareness did not differ significantly amongst participants in the latter three categories (smallest *p* = 0.29). Finally, we found no evidence that either prior discussion or confidence differed as a function of decision categories (smallest *p* = 0.49).

**Figure 6.**
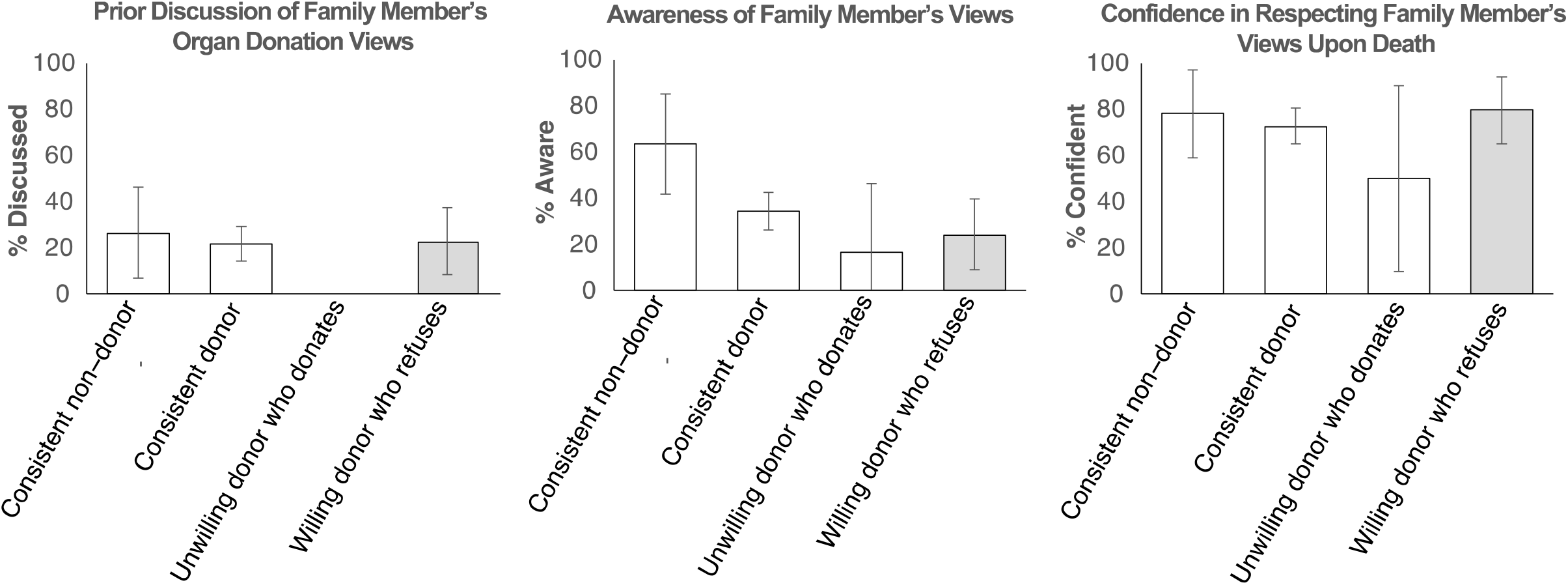
Bar graphs depict the percent of participants in each category (consistent non-donors, consistent donors, unwilling donors who donate relatives’ organs, or willing donors who refuse family donations) who: had discussed organ donation with a close family member (left panel), were aware of their relative’s wishes (middle panel), and were confident of carrying these wishes out (right panel). Vertical lines represent 95% confidence intervals.

## Discussion

Increasingly, social science research underscores how the decision-making process differs when one decides for another person as compared to him-or herself.^37,38^ Through our research, we applied this framework to organ donation in a novel manner, seeking to understand family refusal by accounting for the decision-maker’s mindset (vis-à-vis deciding for the self).

First, analogous to healthcare surrogacy, we observed for the first time how individuals were overconfident in making donation decisions for close family members. Although they had not discussed their family members’ wishes nor were aware of these, participants were nonetheless confident that they could carry out their relatives’ wishes upon death. Likewise, when individuals considered decisions made on their behalf, they expressed confidence that family members would do their bidding – despite not having made their wishes known previously. In both cases, being assured of the family decision-making process far exceeded the basis for doing so.

Our findings of overconfidence provide a baseline for campaigns that urge individuals to discuss organ donation with their families.^39^ Although it is widely recognised that discussion increases family consent, promoting discussion can be challenging, and studies have sought to identify persons most willing to take this step.^40-42^ Beyond person-based characteristics, however, it seems likely that overconfident individuals may find it redundant to initiate discussion, whether to make their wishes known or to find out about their relatives’ wishes. If this account is true, donation campaigns could benefit from addressing overconfidence as a precursor to stimulating family discussion.

In line with previous findings, we further documented a systematic shift when participants made decisions for family members.^18,19^ Namely, participants were less willing to donate their relatives’ organs than their own, with 1 in 5 switching from being willing donors themselves to refusing donation for their relatives. Extending prior research, we also identified for the first time two key factors – fears about organ donation and religion – that predicted when decisions for the family matched decisions for the self.

Examining the two key factors more closely, we found that consistent donors and non-donors were identified by strong influences – having extreme levels of organ donation fears (e.g., low fears predicting consistent donation), or strong religious views (e.g., affiliations with Taoism, Islam, Hinduism, and Catholicism predicting refusal). While each factor had previously been linked to individual and family decision-making independently,^18,43,44^ our findings additionally highlight the role of these factors in decision consistency – making the same organ donation choice for both the self and for a family member. By contrast, the majority of those who switched – namely, willing donors who refused family donation – were less driven by these factors. Instead of relying on strong principles, this group may have relied on heuristics – rule of thumb guidelines that simplify decision-making.^45^ Thus, being unaware of their relatives’ wishes but overconfident in their decision-making abilities, they may have reverted to the conservative decision of refusing organ donation.

If our account of family refusal is true, then decision-making research would suggest that refusal may be minimized if decision-makers can be nudged to avoid heuristics.^45^ Since family refusal rates exceed those of individuals, moving closer to individuals’ actual wishes is likely to increase the supply of transplantable organs. To this end, donation programs may benefit from focusing on decision-makers willing to donate their own organs, urging them to: (1) become aware of their relatives’ wishes (prior to his/her death); or (2) undergo a more thorough decision-making process to work out their relatives’ wishes (instead of simply relying on the conservative heuristic to abstain from organ donation).

## Limitations

We note several limitations of our study. First, to compare decisions for the self versus a family member, we relied on hypothetical willingness to donate. This survey methodology – while commonly used – stands in contrast to retrospective audits of hospital records where actual donation decisions are examined.^6^ Future research will need to determine whether our findings generalize to real-life contexts where – for example – decisions have to be made while standing at the hospital bed of a family member. In a similar vein, the self-reported nature of our study predictors (e.g., whether participants had discussed organ donation, organ donation fears) may have been influenced by recall biases. Finally, although our sample was representative of the general population, all data were collected in an urban setting and may not apply to rural contexts.

## Conclusions

In summary, we conducted an in-depth analysis of psychological processes that may underlie family refusal. Our findings underscore how decision-making differs when one considers a family member rather than the self. Consequently, addressing the decision-maker’s mindset (e.g., overconfidence, the use of heuristics) may increase actualization rates amidst a worldwide organ shortage.

## Data Availability

Due to IRB requirements, data will be provided upon request.

## Acknowledgements

The authors gratefully acknowledge Madhumitha Ayyappan, Rayner Ng, Joanna Chue, Keith Tong, Joel Chew, Chow Kit Ying, Sean Nicholas, Hans Toby Limanto, Sophie Ang, Dinh Hai Bao Lien, Claris Nghai, Pei Jia Ying, and Nasir Ruslan for their assistance in the preparation of surveys, data collection, and data entry.

## Contributors

CWL, LNC, AA, and JCJL participated in research design, data analysis, and the writing of the paper. BLZ, CKYL, and WHN participated in research design and performance of the research. TS and VKH participated in research design and the writing of the paper.

## Data sharing

Data are available upon request (subject to approval from the relevant ethics boards).

